# Modelling the effectiveness of attractive targeted sugar baits in reducing clinical malaria

**DOI:** 10.64898/2026.07.28.26359124

**Authors:** Lars Kamber, Aurélien Cavelan, Emma Louise Fairbanks, Julian Entwistle, Melissa Penny, Nakul Chitnis

## Abstract

Attractive targeted sugar baits (ATSBs) are a novel vector intervention targeting the sugar feeding behaviour of mosquitos by providing toxic sugar meals. ATSBs have shown strong efficacy in terms of mosquito population reduction in entomological trials but their epidemiological impact remains incompletely understood. This modelling study explores the impact of ATSBs on entomological and epidemiological outcomes over a wide range of transmission settings. We quantify the required killing probabilities to achieve target incidence reductions and demonstrate the sensitivity of ATSB efficacy to mosquito characteristics such as sugar feeding frequency and sac rate. Furthermore, we show that ATSBs can maintain reduced incidence levels previously achieved by long-term insecticide-treated net campaigns.

Despite mixed results from clinical trials, these results highlight the high potential of ATSBs as a complementary vector control strategy, while underscoring the critical importance of entomological parameters in predicting public health impact. Realizing this potential will require accurate characterization of mosquito behavior and optimized deployment strategies to ensure sufficient population-level killing probabilities of mosquitoes.

## 1 Background

While great progress in the fight against malaria has been made over past decades, reduction in malaria case burden is stagnating in many locations [44]. Conventional vector control interventions such as insecticide-treated nets (ITNs) and indoor residual spraying (IRS) have been responsible for the majority of these gains, but cannot target residual transmission from outdoor and evening biting mosquitoes [6]. Their effectiveness is further reduced by new challenges, including mosquito behavioural adaptations and growing insecticide resistance. Additional complementary interventions are needed to maintain current achievements, further reduce disease burden, and potentially interrupt transmission in endemic regions.

Attractive targeted sugar baits (ATSBs) represent a promising vector control approach that exploits mosquitoes’ sugar-feeding behaviour [4]. Sugar-feeding in female mosquitoes has been well-documented through methods such as cold anthrone tests that can detect undigested sugar in mosquito midguts [42] and capturing mosquitoes stained with dyed non-toxic sugar solutions, which are also referred to as attractive sugar baits (ASBs) [16, 32, 33, 42]. ATSBs leverage sugar-feeding by combining attractive sugars with insecticides, effectively creating toxic sugar meals that can target both male and female mosquitoes indoors or outdoors - addressing critical gaps in protection not covered by conventional tools. They can be deployed either as solutions sprayed onto vegetation or as bait stations specifically designed to prevent non-target insects and other animals from unintended insecticide exposure. Compared to existing vector control methods, ATSBs offer distinct operational advantages: they eliminate the need for labor-intensive insecticide spraying, larviciding of difficult-to-locate breeding sites, and user adherence with ITN usage; they produce no emissions like repellents and avoid side-effects associated with systemic antiparasitic drugs such as ivermectin.

The hypothesised impact of ATSBs on malaria transmission is two-fold: they directly reduce mosquito population densities and decrease the average mosquito lifespan, which reduces the probability of infected mosquitoes surviving the extrinsic incubation period required for parasite development and onward transmission [42]. Entomological field trials have demonstrated ATSBs efficacy in drastically reducing mosquito populations across various species [5, 16, 25, 26, 32, 33, 34, 36, 37, 42], with some studies reporting population reductions exceeding 90% [24, 34]. Although denser vegetation might potentially reduce ATSB effectiveness by providing competing natural sugar sources [27], it has been demonstrated that ATSBs can maintain strong effects on mosquito populations even in a non-arid setting with more abundant vegetation [32].

An essential quantity for mathematical modelling of the impact of ATSBs on mosquito populations and consequent epidemiological outcomes is the daily feeding rate of mosquitoes on ATSBs. This daily feeding rate is typically translated to an equivalent daily mortality rate [14, 21, 22], as mosquitoes that feed on ATSBs have been shown to die with very high probability, though not with 100% [5, 28]. Based on entomological trial data, this feeding rate has been estimated to be as high as 0.5 per day in a single-site trial in Mali [21, 23], between 0.07–0.11 per day in a cluster-randomised entomological trial in Mali [14, 42], and between 0.04 and 0.09 per day in non-arid western Zambia. Mathematical models incorporating the impact of ATSBs on malaria using these feeding rates predicted substantial reductions in malaria incidence following ATSB deployment [14, 21], potentially outperforming existing vector control interventions and even suggesting the potential for transmission interruption in some scenarios [21].

Building on the promising entomological results and model predictions, Westham Co. developed the Sarabi Bait Station, an ATSB product designed for mass production, long-term deployment, and safe delivery [45]. The efficacy of this product on epidemiological and entomological endpoints was evaluated through three coordinated cluster-randomised trials conducted in Mali, Kenya and Zambia [12, 45]. Prior to these clinical trials, mathematical modelling was conducted to determine the minimum daily ATSB feeding rate required to achieve a 30% reduction in clinical malaria cases in children [14] — the primary endpoint for which the studies were statistically powered to detect. The required feeding rates estimated by the mathematical models were well below ATSB feeding rates estimated from earlier entomological trials [21, 42]. Based on these estimates and a dyeing study conducted in Mali which showed more than 25% of captured mosquitoes being dyed when installing two ASBs per household structure [11], two ATSBs were installed in each eligible household structure within intervention clusters. None of the trials found a significant reduction in malaria incidence or mosquito density [2, 29, 35]. However, a subanalysis from the trial in Mali showed significant effects on malaria incidence in areas with high ATSB coverage and well-maintained baits.

While these trials provide initial evidence about ATSB effects on clinical malaria, mathematical modelling can offer insights about potential effects of ATSBs across a broader range of settings and address additional questions. Using OpenMalaria, a complex agent-based simulator of malaria transmission and control, we model reductions in mosquito density and incidence of clinical malaria across a wide range of transmission settings. We predict the ATSB killing probability for female mosquitoes required to achieve a 30% reduction in clinical malaria incidence in children under various transmission intensities. We explore how mosquito population characteristics — specifically sugar feeding frequency and sac rate (the proportion of parous mosquitoes that have laid eggs within twenty-four hours) — influence these results. Beyond this, we evaluate the potential of ATSBs to maintain reduced malaria incidence levels previously achieved under ITN coverage. Finally, we assess the importance of explicitly modelling the presence of existing vector interventions when studying the impact of ATSBs by comparing the model predictions with and without explicit inclusion of ITNs.

## 2 Methods

We extend a malaria transmission model, OpenMalaria, to include the effects of ATSBs. A simplified schematic of the mosquito model in OpenMalaria including the mechanism of action for ATSBs is provided in Figure 1. OpenMalaria is a stochastic individual-based model designed to analyze malaria transmission dynamics, intervention strategies, and their impacts on public health [1, 9, 10, 38, 39]. It has been used to predict potential outcomes for a wide range of interventions such as insecticide-treated nets [8], mass drug administration [7], and vaccination programs [30]. In humans, OpenMalaria models the asexual and sexual bloodstages of parasites, the development of immunity through infections and interventions, pharmacokinetics and pharmacodynamics, and drug resistance. Severe and non-severe malaria episodes are generated by the disease model, with the treatment pathway for individuals determined by the health system, which can be finely parameterised. The mosquito model in OpenMalaria simulates the life cycle and behaviour of malaria-transmitting Anopheles mosquitoes. The model accounts for essential factors such as feeding behaviour, lifespan, and susceptibility to interventions such as insecticides.

**Figure 1:**
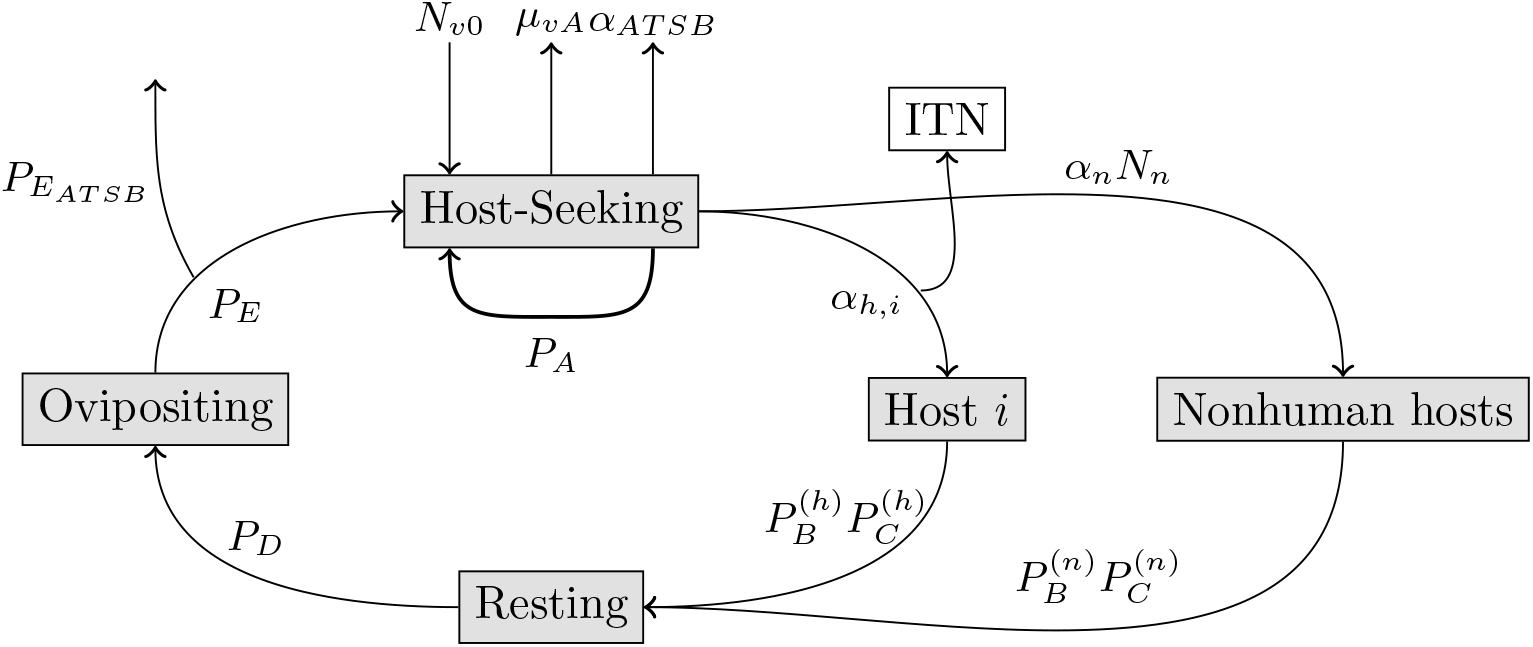
Simplified schematic of the mosquito feeding cycle in OpenMalaria showing ATSB and ITN interventions. Parameters are detailed in Table 1. The model uses daily time steps and models only female mosquitoes. New host-seeking mosquitoes emerge at a rate *N*_*v*0_ which varies under seasonality, creating fluctuating adult female populations. Mosquitoes leave the host-seeking state by: biting human host *i* (rate *α*_*h,i*_), biting nonhuman hosts (rate *α*_*n*_*N*_*n*_), death by ATSB killing (rate *µ*_ATSB_), or natural death (rate *µ*_*vA*_). As a result of these leave rates, mosquitoes leave the host-seeking state with probability 1 *− P*_*A*_ and continue host-seeking the next day with probability *P*_*A*_. After successful host biting, mosquitoes enter a two-day resting/ovipositioning phase with potential mortality due to natural causes or interventions, represented by progression probabilities *P*_*B*_ to *P*_*E*_. ATSB-caused mortality during resting/ovipositing states is captured by 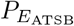. The model explicitly tracks the total number of host-seeking mosquitoes, oocyst-positive (infected) host-seeking mosquitoes and sporozoite-positive (infectious) host-seeking mosquitoes, while implicitly modelling other states.

In the model extension the effects of ATSBs are parameterised as a daily *probability of death* of mosquitoes. As the proportion of mosquitoes that die after feeding on an ATSBs is high but may not be perfect, we define the effect of ATSBs in terms of a *killing* probability and not a *feeding* probability. Internally, OpenMalaria models mosquitoes exiting the host-seeking state using rates, either due to finding a host or due to mortality. The daily probability of death due to ATSBs on a host-seeking day, 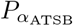, is a function of the rate of death due to ATSBs, *α*_ATSB_(*t*), and all the other rates, with which mosquitoes leave the host-seeking state (see caption Figure 1):

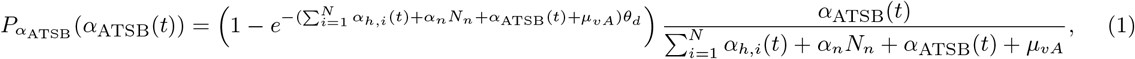

with the parameters given in Table 1. The first term of equation (1) corresponds to the probability that a mosquito leaves the host-seeking state at all, while the second term determines the fraction of mosquitoes that leave the host-seeking state due to feeding on an ATSB. In order to achieve a target daily killing probability, we numerically solve this equation for the required daily death rate, *α*_*AT SB*_(*t*). Because the availability rate of individuals 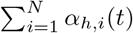 changes over time due to changes in population structure and other factors, such as effective ITN coverage, *α*_ATSB_(*t*) is recalculated every time step to keep *P*_ATSB_ constant.

**Table 1:** Description of parameters with dimensions provided in parentheses, where appropriate.

|  |  |
| --- | --- |
| $P_{\alpha_{\text{ATSB}}}$: | Daily mosquito mortality probability due to ATSBs on host-seeking days |
| $\alpha_{\text{ATSB}}(t)$: | Daily mosquito mortality rate due to ATSBs on host-seeking days calculated for target $P_{\alpha_{\text{ATSB}}}$ (time<sup>-1</sup>) |
| $P_{E_{\text{ATSB}}}$: | Mosquito mortality probability due to ATSBs during the resting and ovipositing stages |
| $\alpha_n$: | Availability rate of individual nonhuman hosts (time<sup>-1</sup>) |
| $\alpha_{h,i}(t)$: | Availability rate of individual $i$ to mosquitoes at time $t$. Depends on age of individual, gamma-distributed biting exposure of individuals and net availability for individuals (time<sup>-1</sup>) |
| $\mu_{vA}$: | Mosquito mortality rate while host-seeking (time<sup>-1</sup>) |
| $\theta_d$: | Proportion of the night that the mosquito spends host-seeking |
| $N$: | Population size of human hosts (humans) |
| $N_n$: | Population size of nonhuman hosts (animals) |
| $P_B$: | Probability of mosquito escaping host after biting |
| $P_C$: | Probability of mosquito entering resting stage |
| $P_D$: | Probability of mosquito entering oviposition stage |
| $P_E$: | Probability of mosquito re-entering host-seeking stage |

We express ATSB effects using the killing *probability* 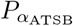 rather than the ATSB killing *rate α*_ATSB_(*t*) because the probability offers more direct interpretation. The impact of a given ATSB killing rate depends on the other rates at which mosquitoes leave the host-seeking state [14]. Maintaining constant killing probability (rather than a rate) prevents unwanted interaction effects with other interventions like ITNs. For example, if we kept the killing rate *α*_ATSB_(*t*) constant while ITNs reduced the rate mosquitoes find hosts 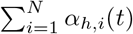, the daily ATSB killing probability would unintentionally increase. Using probability as our parameter avoids this issue, as killing rates can be dynamically adjusted over time to achieve some target killing probability.

Mosquito mortality due to ATSBs on non-host-seeking days is introduced through probability 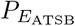. This probability reduces the proportion of mosquitoes returning to the host-seeking state after ovipositing. We assume that after feeding, it takes two days for mosquitoes to oviposit and return to host-seeking. If the probability of ATSB feeding on a non-host-seeking day is *P*_ATSB_, if we assume mosquitoes only feed after ovipositing we set 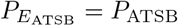 whereas if we assume that mosquitoes feed on days when they rest after they oviposit, we set 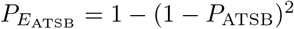.

We assume that heterogeneity in the availability of individuals to mosquitoes (and therefore in the rates at which individuals receive bites *α*_*h,i*_(*t*)) follows a gamma distribution with a coefficient of variation of 1.5 [17]. The availability to mosquitoes is also modulated by age with children receiving fewer bites. We use the same age distribution, based on Tanzanian data, that was used to fit the parameters of OpenMalaria [31]. The effective coverage for uncomplicated malaria cases is 37% [15]. We use a transmission seasonality profile derived from mosquito catchment numbers from Siaya in western Kenya [41]. Based on the same data set, we assume the mosquito species to be *An. funestus*. When modelling the effect of ITNs, we assume 90% of bites occur indoors, a human blood index of 0.98, and breeding sites are the limiting factor in mosquito emergence. The last assumption implies that a change in the number of adult mosquitoes will not result in a change in the emergence rate of mosquitoes. Apart from the study simulations where we explicitly model ITNs, we assume that other interventions, such as ITNs and IRS, are captured implicitly through a reduced EIR. This removes the necessity to explicitly model the present vector interventions, which is computationally expensive, difficult to parameterise due to the frequent lack of historic data on interventions, and makes it harder to generalise the results. We confirm the validity of this approach to implicitly model additional interventions for ITNs (see below). In the absence of clear data, we assume no seasonality in ATSB killing probabilities which may occur due to seasonality in the availability of natural sugar sources. As we directly work with the daily mortality due to ATSBs, we do not consider the spatial placement of ATSBs.

We let the model run for a burn-in period of 115 years in order to ensure stable immunity in the human population when the interventions are first placed. We deploy interventions at the beginning of the transmission season assuming that there is no decay in the effectiveness of ATSBs. We then measure different epidemiological outcomes over 1 year with and without ATSBs with various killing probabilities. For clinical incidence, we measure all cases that occur in one year. For prevalence and mosquito abundance, we compare the point measurements at the time the intervention was deployed compared to the point measurements one year later.

To represent a wide range of transmission intensities for the predicted impact of ATSBs on malaria, we performed a total of approximately 50000 simulations over EIRs between 0.5 and 400. For purely entomological analyses, we run simulations with daily ATSB killing probabilities between 0% and 80%. When considering epidemiological outcomes, we restrict the simulated daily ATSBs killing probabilities to a maximum of 20% despite the higher estimated feeding probabilities in some field experiments [14, 21], because the incidence predicted by OpenMalaria is already reduced to almost zero at killing probabilities above 20% for the considered transmission intensities. To determine the required killing probabilities for a 30% reduction over a range of baseline EIRs and prevalences, we fit a Gaussian process with either one of these variables and the ATSB killing probabilities as the independent variable and the resulting reduction in incidence during the 1-year observation period as the dependent variable using the Python package BoTorch [3]. In order to account for heteroskedasticity, we fit a second Gaussian process on the residuals resulting from the predictions error terms of the predictions of the first Gaussian process. Using these emulator models, we determine the required killing probability for each studied baseline EIR and prevalence using root-finding algorithms.

As a baseline, we assume that mosquitoes only feed on sugar, and therefore, potentially on ATSBs, while host seeking. In this case, mosquito mortality is only introduced through 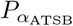 while 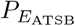 is set to zero. When studying the impact of assuming additional sugar feeding while ovipositing or every day, we set 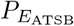 such that it results in the same daily probability of death due to ATSBs on sugar-feeding days as during host seeking.

## 3 Results

### 3.1 Impact of ATSBs on mosquito populations

The predicted relative reduction in mosquitoes for a given ATSB killing probability is independent of the studied EIR setting and, therefore, of the number of mosquitoes initially present. For example, a 1% daily killing probability reduces the mosquito population by 3.4% and a 5% daily killing probability reduces it by 15.2% (Figure 2A).

**Figure 2:**
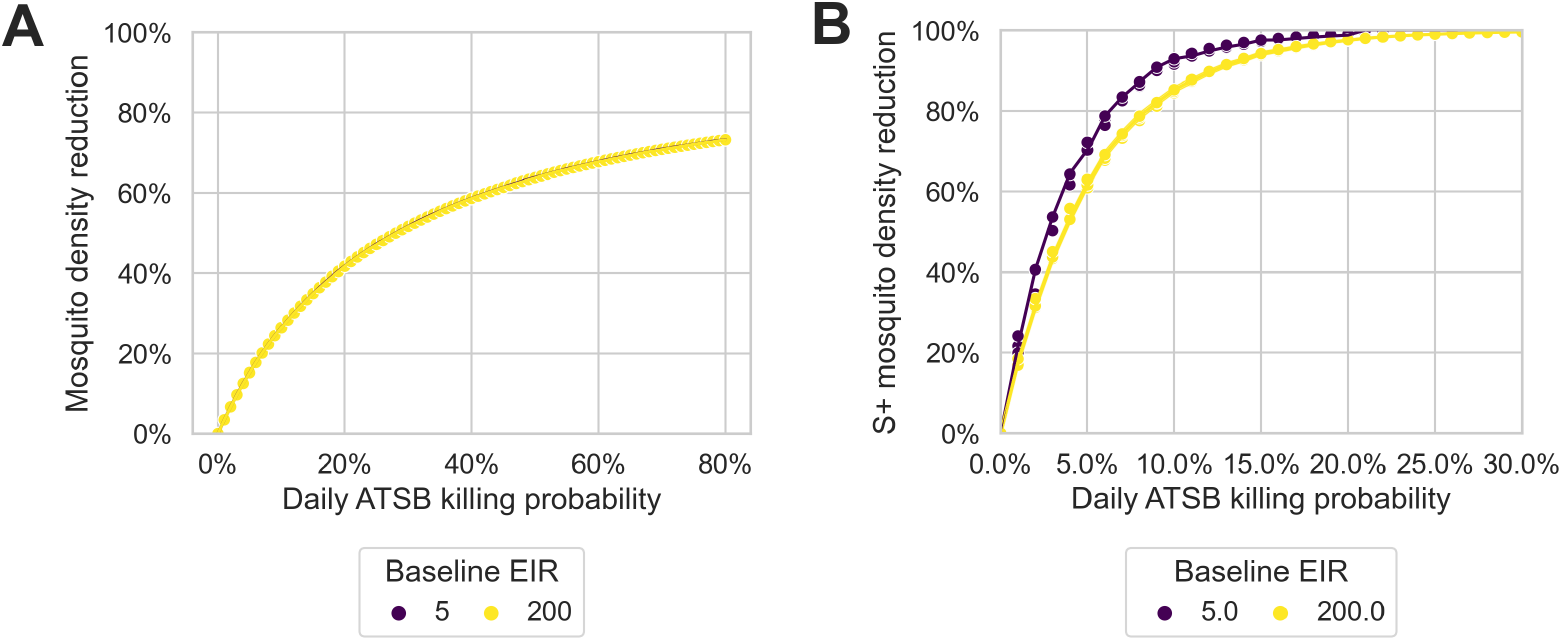
Relative reduction of the overall mosquito population (A) and the population of sporozoite positive mosquitoes (B) over daily ATSB killing probabilities for baseline EIRs of 5 and 200.

The reduction in sporozoite-positive mosquitoes is stronger than the reduction in mosquito density (Figure 2B). Unlike mosquito density, there is a dependence of the relative reduction in sporozoite-positive mosquitoes on the EIR of the setting: assuming a daily 1% ATSB killing probability, the predicted reduction in sporozoite-positive mosquitoes is 21.8% at an EIR of 5, while this reduction is 4% lower at 17.8% at an EIR of 200. Plotting the reduction in mosquito density against the reduction in density of sporozoite positive mosquitoes highlights the disproportionally high reduction in sporozoite-positive mosquitoes by ATSBs (Figure 3). For example, a reduction in mosquito density of 20% results in a reduction in sporozoite-positive mosquitoes of 83% for an EIR of 5 and 74% for an EIR of 200. Comparing the model outputs with the HLC catches from the Mali field data, in the four months with the highest transmission (Jul-Oct), suggests that the model approximates this relationship for reduction in the number of mosquitoes observed in this study [42].

**Figure 3:**
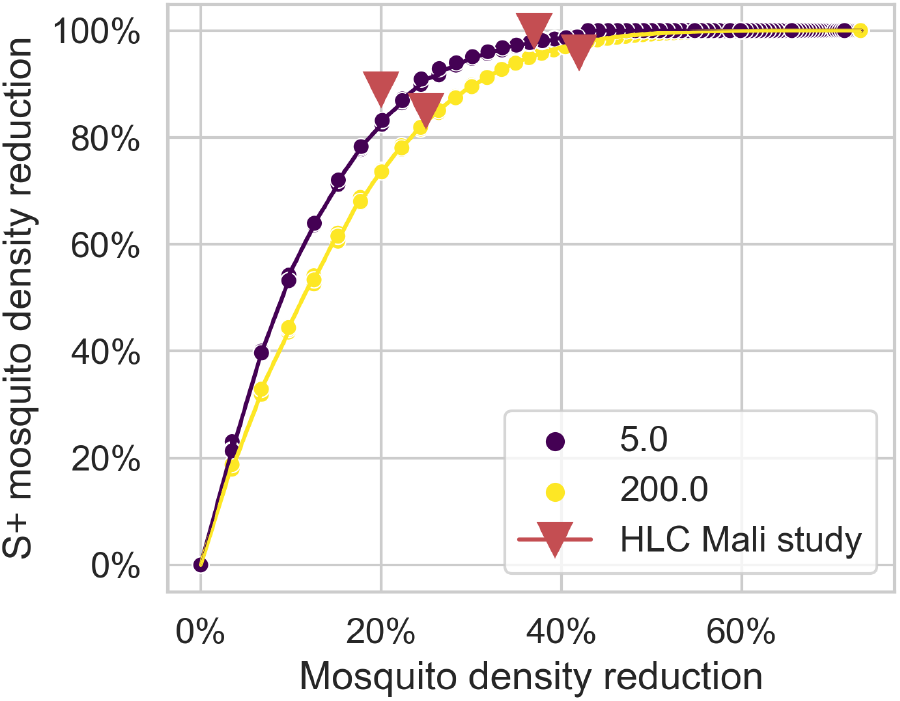
Relative reduction in sporozoite positive mosquitoes over relative reduction in mosquito density achieved by ATSBs. The triangles show data points from the months with highest transmission gathered in an entomological study in Mali [42].

### 3.2 Impact of ATSBs on malaria incidence

The model predicts ATSBs to be highly effective at reducing malaria incidence in children aged 1–14 with the strongest relative reduction in settings with low EIRs (Figure 4A) and, correspondingly, in settings with low prevalence (Figure 4B). For example, at an EIR of 5, the predicted reduction is 11% with a daily killing probability of 1% and 46% with a daily killing probability of 5%. At an EIR of 200, the predicted reduction is 3% with a daily killing probability of 1% and 18% with a daily killing probability of 5%. While the largest relative reductions in malaria incidence are achieved in the lowest transmission settings for all daily killing probabilities, the largest absolute reductions are expected to occur at an EIR of around 10 for killing probabilities of up to 5%. At an EIR of 10, the absolute incidence reduction is 0.13 cases per person per year for a killing probability of 1% and 0.55 cases per person per year for a killing probability of 5% (Figure 4C). Even though the peak in terms of absolute incidence reduction is for killing probabilities up to 5% lies at an EIR of 10, effect of ATSBs in absolute terms is flat over a wide range of prevalences for these killing probabilities (Figure 4D).

**Figure 4:**
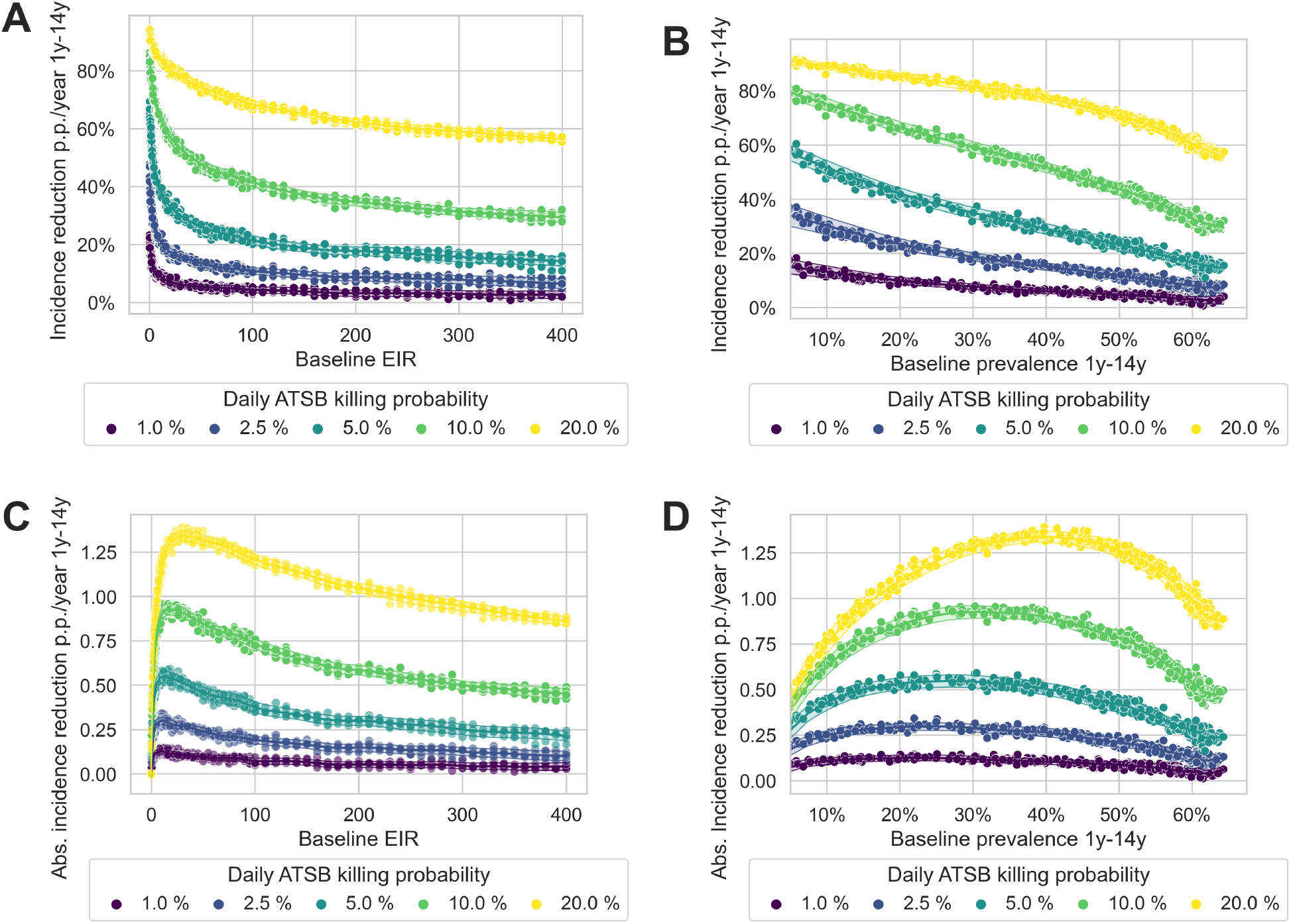
Reduction in incidence of clinical malaria in children aged 1–14 years by ATSBs with various daily ATSB killing probabilities over baseline EIRs (A, C) and baseline prevalences (B, D) before the deployment of ATSBs. The top row (A, B) shows relative incidence reductions, while the bottom row (C, D) shows absolute incidence reductions. Each point represents a single simulation result, the statistical trend and the stochastic uncertainty from the model are shown with lines and shaded areas.

The required killing probability to achieve a 30% reduction in malaria incidence is estimated to be 2.8% for an EIR of 5 and 8.4% for an EIR of 200 (Figure 5A). To achieve a reduction of 70% in malaria incidence, the required killing probabilities are substantially higher: A 9.4% killing probability is required at an EIR of 5 and a 24.5% probability is required at an EIR of 200. The relationship is concave of the required killing probabilities and is concave over increasing EIR, while it is convex over increasing prevalences (Figure 5B). This is a consequence of the EIR-prevalence relationship, as simulated by OpenMalaria, where prevalence increases faster with increasing EIR than the required ATSB killing probabilities.

**Figure 5:**
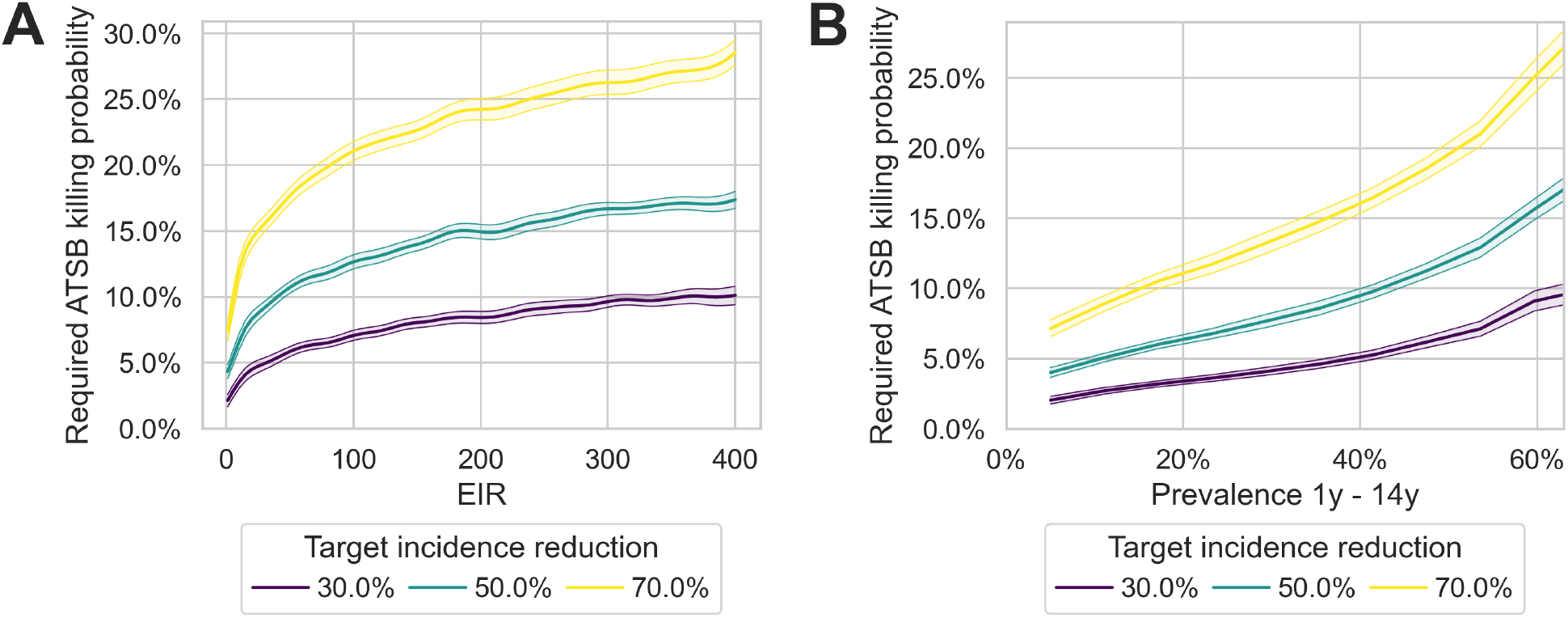
Required daily killing probabilities of mosquitoes due to ATBSs ATSBs to achieve a 30%, 50% or 70% reduction in clinical malaria incidence in children aged 1-14 years over in the first year after deployment. Subfigure A shows the required killing probabilities over a range of baseline EIRs, subfigure B over a range of baseline prevalences.

The reduction in incidence is disproportionately greater than the reduction in mosquito density caused by ATSBs (Figure 6). The incidence reduction is almost linear in the mosquito density reduction up to a point of saturation, where further incidence reductions are not possible as disease transmission is reduced to almost zero, The effect of reduced mosquito density on incidence reduction is more disproportionate with a lower baseline EIR: At a baseline EIR of 5, a reduction of mosquito density of 20% results in an incidence reduction of 59% while the same reduction in mosquito density under a baseline EIR of 150 only results in a 27% reduction in incidence.

**Figure 6:**
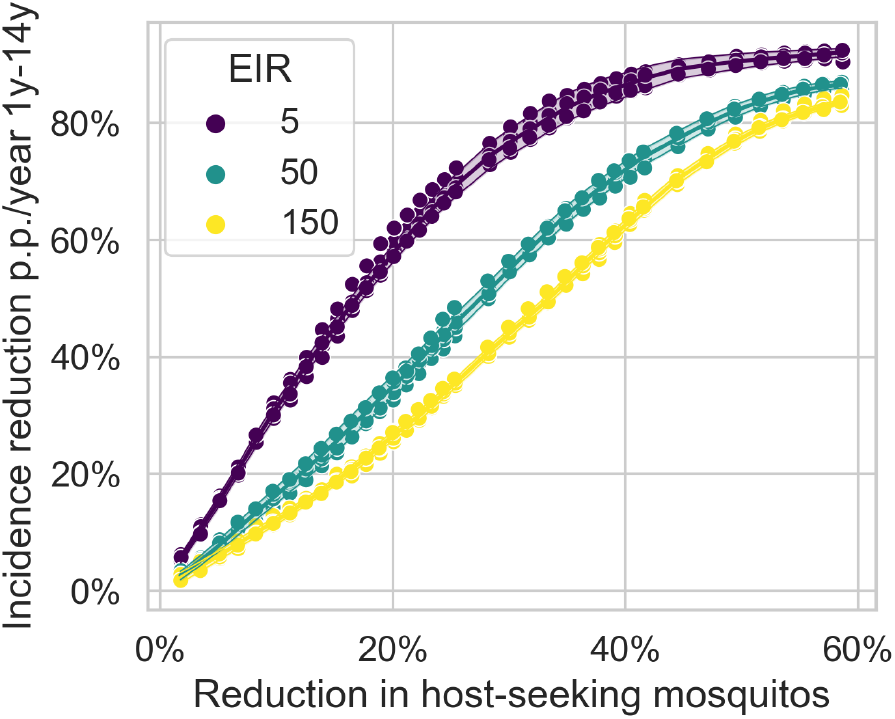
Reductions in mosquitoes caused by ATSBs and the resulting reduction in incidence in children aged 1 – 14 years for EIRs 5, 50, and 150.

### 3.3 Impact of sugar feeding frequency assumptions on ATSB efficacy

The assumptions regarding when mosquitoes seek sugar, and therefore feed on ATSBs, substantially affect predicted reductions in mosquito density and malaria incidence. When mosquitoes are assumed to feed on ATSBs only during host-seeking, rather than also during resting and ovipositing, the efficacy of ATSBs is significantly reduced. At a baseline EIR of 50, a daily 2% ATSB killing probability reduces the mosquito density by only 7% when sugar feeding is limited to the host-seeking stage, compared to a 12% reduction during all stages of the mosquito feeding cycle (Figure 7A). Consequentially, the required daily killing probabilities to achieve a 30% reduction in malaria incidence are almost twice as high when assuming sugar-feeding only during host-seeking, across the entire range of transmission intensities (Figure 7B). For example, at a baseline prevalence of 40%, the required killing probability increases from 1.9% when assuming sugar feeding all stages of the mosquito feeding cycle to 3.8% when assuming sugar feeding only during host seeking.

**Figure 7:**
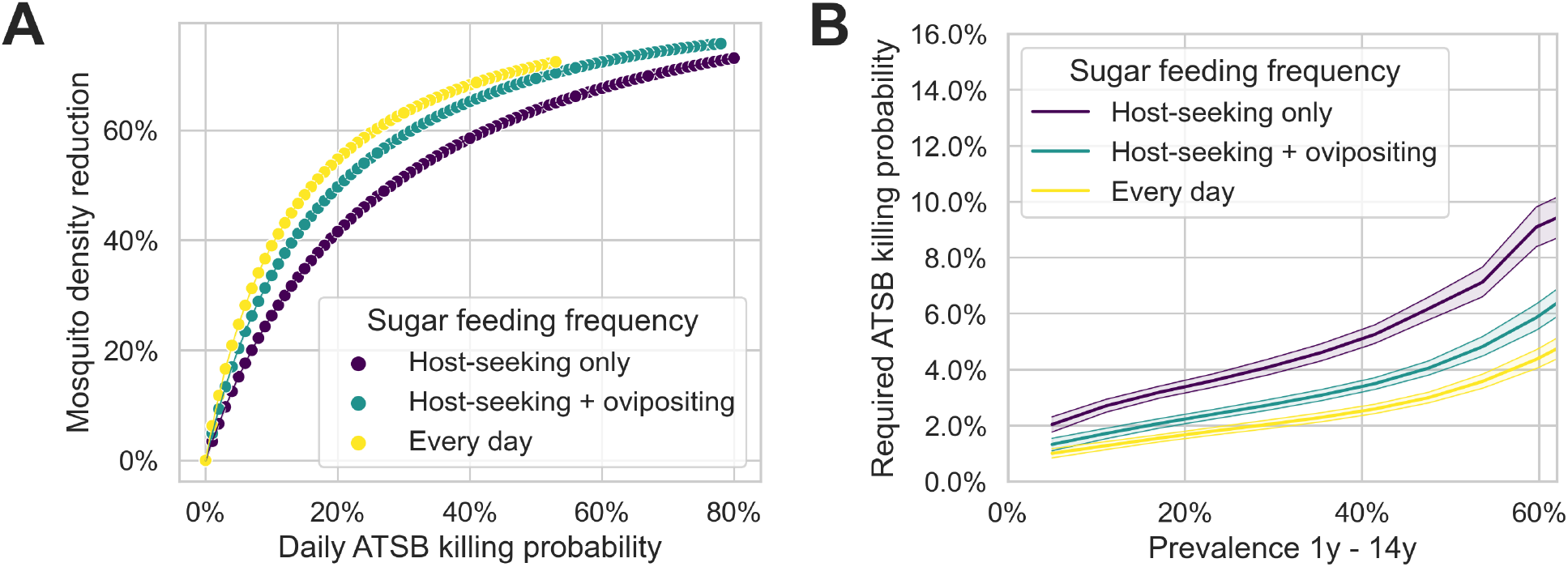
Effects of the ATSB feeding frequency assumption on mosquito density reduction (A) and the required daily ATSB killing probabilities to achieve a 30% reduction in malaria incidence across different baseline prevalences (B).

The sac rate — defined as the proportion of mosquitoes that laid eggs within the last 24 hours — further modulates the impact of ATSBs depending on the feeding frequency assumption. A higher sac-rate implies that mosquitoes are quicker to find a blood meal, consequentially spending less time in the host-seeking state. Accordingly, when assuming that mosquitoes only sugar-feed during host-seeking, a higher sac rate indicates less exposure to ATSBs and therefore leads to smaller reductions in mosquito density and malaria incidence (Figure 8).

**Figure 8:**
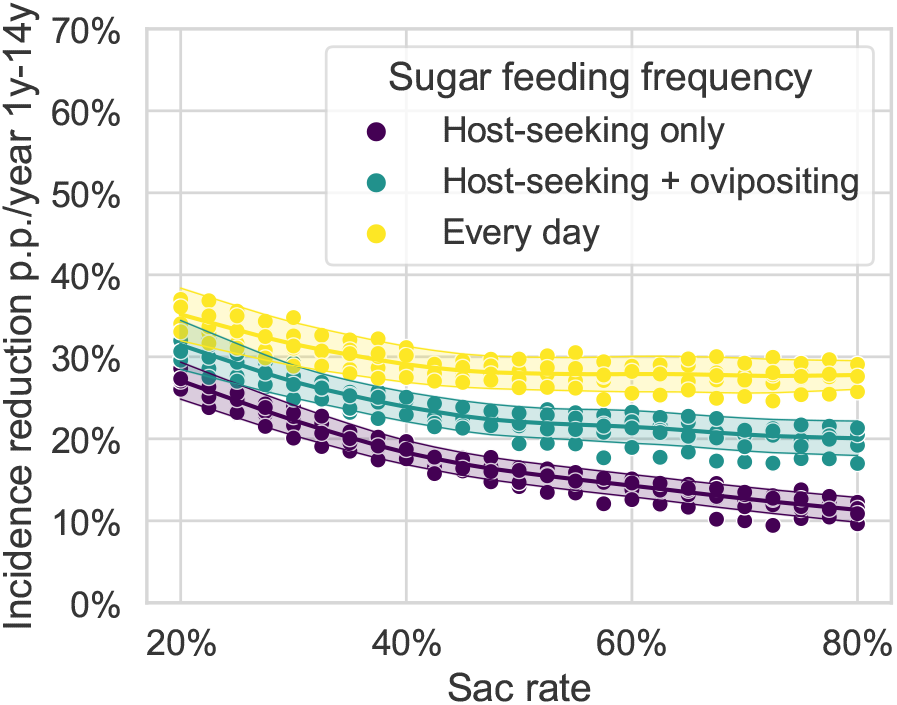
Interaction of sugar-feeding frequency and sac-rates at an EIR of 50 and an ATSB killing probability of 2.5% on days on which mosquitoes feed on sugar.

### 3.4 Interactions of ATSBs with ITNs

We do not find an impact of explicitly modelling the presence of ITNs when ATSBs are deployed. For any given prevalence that was achieved under 15 years of repeated ITN distribution in the model or without the presence of such an intervention, the effect of ATSBs on malaria incidence is the same. Figure 9 shows the complete overlap of malaria incidence reduction with these two scenarios when assuming an ATSB killing probability of 2.5%. This result shows that explicit modelling of already present ITNs does not affect our predicted impact of ATSBs.

**Figure 9:**
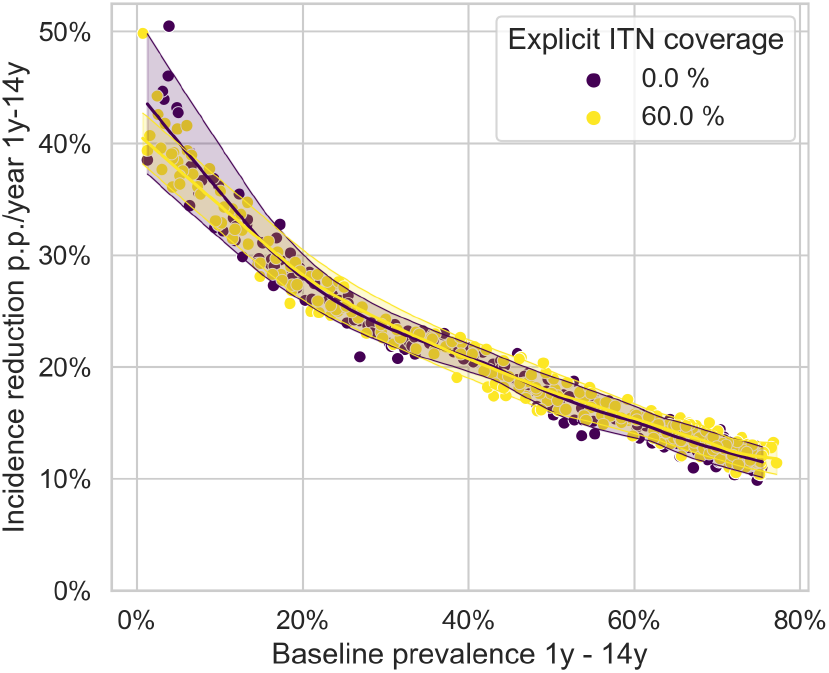
Malaria incidence reductions achieved with a 2.5% daily killing probability for baseline prevalences that were either achieved with an explicit ITN coverage of 0% or an explicit ITN coverage of 60%.

The daily killing probabilities of ATSBs required to sustain incidence reductions while replacing ITNs does not depend on the transmission intensity achieved in the ITN campaign (Figure 10). For example, independently of whether an ITN campaign with 80% coverage resulted in a 20% or 50% prevalence, the same ATSB killing probability of 4.1% (3.9%-4.4%) is required to maintain the level of transmission achieved under the ITN campaign. This result is not contradictory to other results, where the impact of ATSBs depends on the baseline prevalence; This flat relationship rather suggests that ATSB deployment with a daily killing probability between 3.9% and 4.4% is equivalent to carrying on an 80% ITN campaign for any transmission setting. Likewise, killing probabilities between 3.0% and 3.8% are required to match the effect of ITNs at a coverage of 60% and killing probabilities between 2.3% between 3.1% are required to match the effect of ITNs at a coverage of 40% .

**Figure 10:**
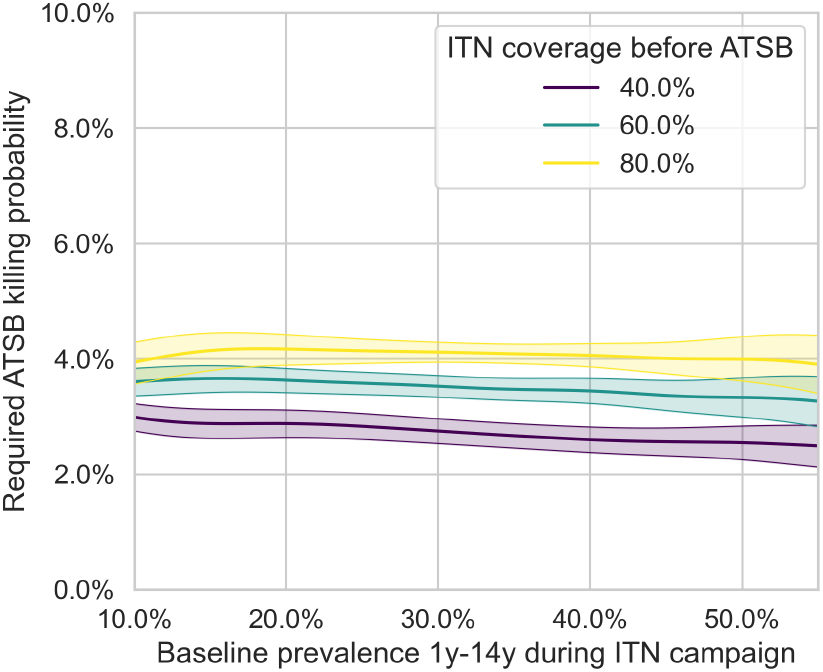
Daily killing probability that ATSBs must achieve to maintain the same level of malaria incidence as ITNs, if ATSBs were deployed in place of ITNs at 15 years of repeated ITN distribution.

## 4 Discussion

Our results confirm previous simulation studies showing a high potential of ATSBs to reduce the incidence of clinical malaria cases [14, 21, 22]. We predict that daily killing probabilities of mosquitoes on ATSBs below 5% can result in a 30% reduction in the incidence of clinical malaria in transmission settings with an EIR of up to 50 infectious bites per person per year. We show the potential of ATSBs to sustain the reductions achieved by the mass distribution of ITNs over the past decade as an alternative to continued net distribution, again at killing probabilities below 5% even in the highest transmission settings. We also demonstrate that the feeding frequency of mosquitoes greatly impacts the effects of ATSBs. Achieving target incidence reductions requires almost twice the daily killing probability if mosquitoes are assumed to only sugar-feed while host seeking, as opposed to sugar-feeding every day. As a further example of the importance of mosquito characteristics, we show that the sac-rate parameter greatly modulates the predicted impact of ATSB with a higher sac rate drastically reducing the effectiveness of ATSBs. Finally, we show that the approach of not explicitly modelling present vector interventions does not affect the predicted impact of ATSBs with our parameterisation.

The apparent contrast between the positive modelling results shown here and the recently published trial results requires explanation given that these trials showed no significant effect of ATSBs on mosquito density and incidence of clinical malaria [2, 29, 35, 43]. The absence of a reduction in the relevant mosquito population transmission corresponds to an ATSB killing probability of 0% in our model, with no predicted effect on malaria transmission. Factors that could have affected the impact of ATSBs on the mosquito population include the ATSB density, the spatial placement of ATSBs, the height of ATSB placement, ATSB maintenance, ATSB design, the attracting ingredients, and the interaction with competing sugar sources or blood meal opportunities.

A potential issue is an incomplete overlap between the mosquito population that drives malaria transmission and the mosquito population that feeds on ATSBs and is trapped for measurement of mosquito population reduction. This may have resulted in the dye experiments, conducted prior to the RCT to inform the required ATSB density, overestimating the proportion of relevant mosquitoes that fed on ASBs with the tested ASB densities. Consequently, the true feeding probabilities of the mosquito population driving malaria transmission could be substantially lower than calculated from the dyeing experiments [22]. In the clinical trials in Zambia, though non-significant, the mosquito population reduction was substantially stronger than the incidence reduction, which would suggest a bias in the mosquito population measurements. Based on our modelling results, we expect a disproportionately higher reduction in malaria incidence for a given reduction in mosquito density if the mosquito population affected by ATSBs and the mosquito population driving transmission are the same: a result consistent with George Macdonald’s analysis showing the disproportionate impact of reducing the daily survival of mosquitoes [19, 20].

The model faces several limitations. While the use of a killing probability instead of a rate makes for easier interpretation and abstracts away from all the complications discussed above, the use of rates might better represent how different interventions interact, especially under changing coverage. The result of explicit ITN coverage modelling that does not affect the impact of ATSBs could also crucially depend on working with constant ATSB killing probabilities instead of rates. However, estimating the killing rates of ATSBs correctly requires knowledge of all other rates with which mosquitoes leave the host-seeking state. In conclusion, either ATSB killing rates or probabilities estimated using field data are likely to be highly setting-specific for the natural environment and interventions affecting the mosquito population where the data was collected. This point is further emphasised by the impact of the sac rate on the predicted reduction in incidence.

The model can currently not distinguish between mosquitoes being outdoors or indoors and therefore, the fact that the ATSBs are only placed outdoors cannot be explicitly be considered. This has also not been the case in previous ATSB modelling studies and is a direction worth exploring, given the possibility to place ATSBs outdoors is often discussed as an advantage complementing the more established indoor interventions such as IRS and ITNs [13, 21].

We have not considered the potential decrease in sugar feeding frequency of aging as mosquitoes progress through gonotrophic cycles [18, 40, 21]. Relatively increasing the sugar feeding frequency in younger mosquitoes would decrease the number of mosquitoes surviving the extrinsic incubation period and therefore further increase the efficacy of ATSBs. Male mosquitoes, which exclusively feed on sugar sources, and therefore have a high potential to feed on ATSBs, are currently not modelled in OpenMalaria. Strong reductions in males could affect the emergence of new mosquitoes. Furthermore, by killing male mosquitoes, ATSBs may affect the evolution of insecticide resistance, which is not considered here. Finally, the model assumes that the emergence rate of new mosquitoes does not change with mosquito density, following the assumption that saturated breeding sites are the limiting factor in the emergence of new mosquitoes. Including the effect of ATSBs on male mosquitoes and density-dependent mosquito emergence would further increase the predicted impact of ATSBs on the mosquito population.

Despite mixed results from initial clinical trials, our modelling demonstrates the significant potential of ATSBs as a complementary vector control tool for reducing malaria transmission. With daily killing probabilities of just 5% or less, ATSBs could achieve meaningful reductions in clinical malaria incidence across various transmission settings, and potentially interrupt transmission in lower-prevalence areas. If such daily killing probabilities are feasible, ATSBs offer a promising alternative mechanism targeting mosquito sugar-feeding behaviour that could be particularly valuable in outdoor settings where ITNs and IRS are less effective, and where mosquitoes continue to develop resistance to conventional interventions. However, these reductions in daily mosquito survival would need to be consistent across all mosquitoes transmitting malaria in the intervention area. Further optimisation of ATSB deployment strategies, informed by the insights from our modelling of mosquito population characteristics and feeding behaviours, may help realise the full potential of this innovative intervention and contribute significantly to malaria control and elimination efforts globally.

## Data Availability

All data produced in the present work are contained in the manuscript.

## Supporting information

Figure S1 shows and example time series of scenarios exploring the effect of replacing ITNs with ATSBs. In all simulations, ITNs are deployed every 3 years from year 10 to year 25 at a coverage of 60%. At year 25, there is either another round of ITN distribution or distribution of ATSBs. We run a range of simulations with different daily killing probabilities on ATSBs to determine the killing probability that results in the same incidence as the continued ITN distribution.

## Acknowledgments

We thank David Malone (Gates Foundation) for the suggestion of considering the interaction of ATSBs and ITNs; and Christen Fornadel, Angela Harris, Derric Nimmo, Jason Richardson and Fred Yeomans from IVCC forreviewing the manuscript and providing valuable input. Calculations were performed at sciCORE (http://scicore.unibas.ch/) scientific computing core facility at University of Basel. Emma Fairbanks acknowledges support from a University of Manchester Healthier Futures Fellowship and L’Oréal-UNESCO For Women in Science Award.

